# Cardiovascular Mortality during the COVID-19 Pandemics in a Large Brazilian City: a Comprehensive Analysis

**DOI:** 10.1101/2021.10.08.21264699

**Authors:** Luisa C C Brant, Pedro C Pinheiro, Antonio L P Ribeiro, Isis E Machado, Paulo R L Correa, Mayara R Santos, Maria de Fatima Marinho de Souza, Deborah C Malta, Valéria M A Passos

**Affiliations:** School of Medicine, Universidade Federal de Minas Gerais, Belo Horizonte, MG, Brazil; Municipal Health Secretariat, Belo Horizonte, MG, Brazil; School of Medicine, Universidade Federal de Ouro Preto, Ouro Preto, MG, Brazil; Vital Strategies, São Paulo, SP, Brazil; Nursing School, Universidade Federal de Minas Gerais, Belo Horizonte, MG, Brazil

**Keywords:** COVID-19, Cardiovascular Diseases, Mortality, Hospitalization, Social Vulnerability

## Abstract

**Introduction:** The impact of COVID-19 pandemics on cardiovascular diseases (CVD) may be caused by health system reorganization and/or collapse, or from changes in the behaviour of individuals. In Brazil, municipalities were empowered to define regulatory measures, potentially resulting in diverse effects on CVD morbimortality.

**Objective:** To analyse the impact of COVID-19 pandemics on CVD outcomes in Belo Horizonte (BH), the 6th greater capital city in Brazil, including: mortality, mortality at home, hospitalizations, intensive care unit utilization, and in-hospital mortality; and the differential effect according to sex, age range, social vulnerability, and pandemic’s phase.

**Methods:** Ecological study analysing data from the Mortality and Hospital Information System of BH residents aged ≥30 years. CVD was defined as in Chapter IX from ICD-10. Social vulnerability was classified by a composite socioeconomic index as high, medium and low. The observed age-standardized rates for epidemiological weeks 10-48, 2020, were compared to the expected rates (mean of 2015-2019). Wilcoxon rank-sum test was used to test differences, and risk ratios with their 95% confidence intervals were calculated. National demographic estimates was used to calculate rates.

**Results:** We found no changes in CVD mortality rates (RiR 1.01, 95%CI 0.96-1.06). However, CVD deaths occurred more at homes (RiR 1.32, 95%CI 1.20-1.46) than in hospitals (RiR 0.89, 95%CI 0.79-0.99), as a result of a substantial decline in hospitalization rates, even though proportional in-hospital deaths increased. The rise in home deaths was greater in older adults and in had an increasing gradient in those more socially vulnerable (RiR 1.45); for high (RiR 1.45), medium (RiR 1.32) and low vulnerability (RiR 1.21).

**Conclusion:** The greater occurrence of CVD deaths at home, in parallel with lower hospitalization rates, suggests that CVD care was disrupted during the COVID-19 pandemics, which more adversely affected older and more socially vulnerable individuals, exacerbating health inequities in BH.

## Introduction

The COVID-19 pandemic had an indirect impact on cardiovascular diseases (CVD) care worldwide, resulting from reorganization of health systems, competing demands, and the change in populations’ behaviour that may have avoided contact with health facilities due to fear of contagion, or adherence to social distancing policies.^1,2^ Previous studies have shown that excess mortality during the pandemic cannot be fully explained by COVID-19 deaths, and in some countries such as the US and the UK, CVD deaths increased in 2020.^3,4^ However, in other countries like Denmark, although more deaths at home were reported in individuals with CVD, overall mortality rates did not increase in the country during 2020.^5^

In Brazil, where nearly 200,000 excess deaths were reported in 2020,^6^ CVD deaths also rose from March to June of 2020, in six capital cities, particularly in the least developed ones, in an analysis that evaluated preliminary mortality data.^7^ However, due to the lack of national coordinated actions, the pandemic affected the country heterogeneously, leading to diverse death rates across the country, which peaked at different periods, and mostly affected socially vulnerable groups.^6,8^ As a response to this lack of national coordination, each municipality had the power to implement local policies to mitigate the pandemic’s impact, including business and school closures, according to the pandemic course in their location.^9^

Belo Horizonte is a city in Brazil with 2.5 million inhabitants, where local authorities daily monitored transmission rates and hospitalizations to guide regulatory measures, and although the city was able to delay the first pandemic wave in comparison to other capital cities – 1877 deaths due to COVID-19 occurred in 2020,^10,11^ also mainly affecting the most vulnerable groups.^9,12^ Moreover, during the 2020 pandemic period, CVD hospitalizations had a 16.3% decline, although the impact on CVD mortality has not been evaluated.^13^

Because the effect of the pandemic on CVD depends on the pandemic course and behavioural changes in a location, a comprehensive analysis of the pandemic’s impact on CVD in a city is fundamental to guide future actions. As such, we aimed to analyse how the COVID-19 pandemic affected CVD deaths in a Brazilian city. To better understand the change in trends we investigated deaths, deaths at home, hospitalizations, ICU admissions and in-hospital mortality, and the differential impact of the pandemic on CVD according to sex, age group, social vulnerability, and the pandemic’s phase.

## Methods

### Study design and Setting

This is an ecological, time-series study conducted in Belo Horizonte city, the 6^th^ largest city in Brazil, with 2.5 million inhabitants, located in the Southeast region.^14^ Brazil has a public universal health system (Unified Health System, Sistema Único de Saúde, SUS), being consultations, hospitalizations, and most medications are free of charge to all Brazilian citizens,^15^ although 50% of Belo Horizonte residents have private health insurance.^16^ During the COVID-19 pandemics, Belo Horizonte’s municipality composed a committee of specialists to address the pandemics issues and help local authorities to decide about the timely implementation of regulatory measures, following three daily indicators: transmission rate, and the proportion of occupied hospital and ICU beds.^9^ Other actions taken by the municipality to confront the pandemic have been published.^12^ We evaluated the effect of the pandemic from epidemiological weeks (EW) 10 to 48, 2020, on CVD outcomes in Belo Horizonte city following the STROBE guidelines for reporting observational studies.^17^

### Data Sources

We analyzed data on the number of deaths and deaths at home from the Mortality Information System (*Sistema de Informação sobre Mortalidade, SIM*, in portuguese). SIM is the official source of mortality data in the country, and is qualified for causes of death before disclosure, which therefore occurs with a 4-months delay. SIM’s quality is rated 4 stars by the “The Global Burden of Disease” study’s star rating system (0-5 stars) to assess the quality of cause of death data.^18^ The numbers of hospital admissions, Intensive Care Unit (ICU) admissions and in-hospital deaths derive from the Hospital Information System (*Sistema de Internações Hospitalares, SIH*, in portuguese), which is also used for administrative purposes, including hospital payments, precluding underreporting. The SIH dataset included all public hospitalizations of the city, from which data for residents of Belo Horizonte were extracted. Previous SIH data quality checks revealed that, on average, after a 3-months period of the hospitalization date, the completeness of data achieves 90% (**Supplementary Material 1**).^9^ Data from both systems were provided by the Belo Horizonte Health Department (*Secretaria Municipal de Saúde, SMS*, in portuguese). SIH data was extracted in March/2021 and SIM data in May/21. As such, the analysis was made including data until epidemiological week (EW) 48/2020. Datasets had census tract information that allowed analysis based on the Health Vulnerability Index (HVI), a social vulnerability index.

HVI is a census tract level health vulnerability index developed by Belo Horizonte Health Department based on sewage system and socioeconomic information (SMS, 2013). The original HVI has four levels Low, Medium, High and Very High, but in the present work High and Very High categories were aggregated and further on named “High”, to avoid small number of cases for specific cause’s analysis.^19^

Because Brazil’s last available census was held in 2010, to estimate rates for 2020 at the municipal level and by HVI we used population estimates that applied growth rates estimated from a population projection developed by Freire *et al* (2019).^20^ Population from census tracts (IBGE, 2011)^14^ with the same HVI was aggregated by age and sex, then age-specific growth rates for a ten-year period (2010 to 2020) estimated for Belo Horizonte was applied to each group of HVI population. More detailed information has been previously published.^9^

### Definition of Causes and Measures

To have a comprehensive understanding of the pandemic’s impact on CVD, we examined outcomes for each of the following causes, by epidemiological week (EW), in adults ≥ 30 years: total CVD, defined as all records within Chapter IX (I00-I99) of the 10^th^ International Classification of Diseases (ICD-10); Heart Failure (ICD-10 I50); Stroke (ICD-10 I60, I61, I62, I63, I64, I678); and Acute Coronary Syndrome (ACS) (ICD-10 I20, I21, I22, I23, I24).^21^ We additionally evaluated all records within Chapter XVIII (R00-R09) to better understand whether changes in the mortality patterns could be explained by changes in ill-defined codes, suggesting poorer quality of death certificates during the pandemics period.

For each CVD cause, seven measures were analyzed: death rates, death at home rates, hospitalization rates, Intensive Care Unit (ICU) admission rates, in-hospital death rates, and proportion of hospital admissions that resulted in ICU utilization or death. The cause of death was classified as the underlying cause, according to the World Health Organization protocols.^22^ Proportions of in-hospital deaths were defined as the ratio between the number registered for each variable and the total number of deaths during hospitalizations, considering each age group, sex and cause. All measures were age-standardized considering as standard the population of Belo Horizonte in 2020. Data for total CVD was then analyzed stratified by sex, age groups (30-59 and ≥ 60 years), and according to vulnerability (high, medium and low HVI).

### Statistical Analysis

For each group (sex, age, cause, measure, vulnerability, and phase), the observed age-standardized rates and proportions for EW 10-48 in 2020 were compared to the expected measure, defined as the 2015 to 2019 corresponding mean. Risk ratios were calculated by dividing the observed by the expected values for each measure. We also calculated confidence intervals for all measures.^23^

To investigate the effect of the pandemic course in the measures above described, we further analysed them according to the pandemic phase in the city. The first confirmed COVID-19 case in the city in fact occurred in EW 9. The first phase comprises EW 10-21 and marks the beginning of the pandemic, when the city adopted its first distancing measures and the number of COVID-19 cases, hospitalizations and deaths were relatively low. The second phase (EW 22-30) starts after the first gradual reopening of business facilities at EW 21, when the peak of COVID-19 occurred in Belo Horizonte in 2020, and the number of adverse events rose sharply. The third (EW 31-48) and last analyzed phase comprises the post-peak period, when the city gradually reopened at EW 31.^24^ All rates were estimated considering the exposure time, allowing comparison between periods. The number of events in each period was divided by the fraction of the population of the corresponding number of weeks of exposure.

For the time series analysis, the five-week moving averages of the studied variables from January to December, 2020, were compared with the five-week moving averages of the previous period (January 2015 to December 2019). In the presented tables, the unsmoothed observed variables were compared to their unsmoothed mean for the same period from the 2015-2019 (EW 10-48). Analyses were performed using R, version 4.0.1.^25^

### Ethical considerations

The study was approved by the Universidade Federal de Minas Gerais (UFMG) and SMS-BH Review Board (Protocol: CAAE 39778720.4.3001.5140). The data that support the findings of this study are publicly available at https://datasus.saude.gov.br/transferencia-de-arquivos/, and the steps to access the data described in a tutorial (**Supplementary Material 2**). However, the information on HVI, which is considered sensitive data, is not available and could only be provided after ethical approval.

## Results

From EW 10-48, 2020, the all-cause age-standardized mortality rates rose from 911 to 1,075 per 100,000 inhabitants in Belo Horizonte. However, **Table 1** reveals no excess mortality in the period for total CVD (RiR 1.0, CI 95% 0.96-1.06), even though there was a displacement in the place of death: while deaths at home increased (RiR 1.32, CI 95% 1.20-1.46), in-hospital mortality rates reduced (RiR 0.86, CI 95% 0.74-0.78), although the proportion of in-hospital mortality increased (RiR 1.16%, CI 95% 1.04-1.30). Looking at specific causes, for ACS and stroke there was no overall excess mortality, but ACS deaths occurred more at home, and were reduced in-hospital. For HF, mortality rates increased 32%, as a result of a significant 53% increase in deaths at home and no significant changes in in-hospital mortality. In parallel with these changes in mortality rates, total CVD hospitalizations rates decreased in the period, as did ICU admission rates, though the proportion of ICU utilization increased. For causes classified within the ICD Chapter XVIII, there was a reduction in mortality during the period, with non-significant changes in hospitalizations’ measures (**Supplementary Material 3**).

**Table 1:** Age-standardized rates per 100,000 inhabitants or proportions for cardiovascular disease outcomes observed in 2020 and expected (mean of 2015-2019) for epidemiological weeks 10-48, their absolute difference, and risk ratio. Belo Horizonte, MG, Brazil.

|  | Observed | Expected | Absolute Difference | Risk Ratio |
| --- | --- | --- | --- | --- |
| **Deaths** |  |  |  |  |
| CVD | 247 (238;256) | 245 (236;254) | 2.0 | 1.01<br>(0.96;1.06) |
| ACS | 41<br>(37;44) | 40<br>(37;44) | 0.2 | 1<br>(0.88;1.14) |
| Stroke | 60 (56;65) | 61<br>(56;65) | -0.5 | 0.99<br>(0.89;1.1) |
| HF | 24 (21;27) | 18<br>(15;20) | 5.9 | 1.32<br>(1.1;1.59)* |
| **Deaths at Home** |  |  |  |  |
| CVD | 84 (78;89) | 63 (59;68) | 20.6 | 1.32 (1.2;1.46)* |
| ACS | 10 (8;12) | 7<br>(6;9) | 2.8 | 1.38 (1.04;1.82)* |
| Stroke | 9<br>(7;11) | 7<br>(6;9) | 1.7 | 1.22 (0.92;1.63) |
| HF | 8<br>(7;10) | 5<br>(4;7) | 3.0 | 1.53 (1.12;2.1)* |
| **Hospital Admissions** |  |  |  |  |
| CVD | 782<br>(766;799) | 1026<br>(1008;1045) | -244.0 | 0.76<br>(0.74;0.78)* |
| ACS | 176<br>(169;184) | 222<br>(214;231) | -45.8 | 0.79<br>(0.75;0.84)* |
| Stroke | 153<br>(146;160) | 148<br>(141;155) | 5.5 | 1.04<br>(0.97;1.11)* |
| HF | 161<br>(154;169) | 168<br>(160;175) | -6.4 | 0.96<br>(0.9;1.02) |
| **ICU Admissions** |  |  |  |  |
| CVD | 299<br>(289;309) | 341<br>(330;352) | -41.6 | 0.88<br>(0.84;0.92)* |
| ACS | 127<br>(121;134) | 155<br>(148;163) | -28.4 | 0.82<br>(0.76;0.88)* |
| Stroke | 48<br>(44;52) | 41<br>(37;45) | 7.4 | 1.18<br>(1.04;1.33)* |
| HF | 45<br>(42;50) | 43<br>(39;47) | 2.8 | 1.06<br>(0.94;1.21) |

| % ICU Admissions |  |  |  |  |
| --- | --- | --- | --- | --- |
| CVD | 38<br>(37;39) | 33<br>(32;34) | 5.0 | 1.15<br>(1.1;1.21)* |
| ACS | 72<br>(70;74) | 70<br>(68;71) | 2.1 | 1.03<br>(0.96;1.1) |
| Stroke | 31<br>(29;33) | 27<br>(25;29) | 3.8 | 1.14<br>(1.01;1.29)* |
| HF | 28<br>(26;30) | 25<br>(23;27) | 2.8 | 1.11<br>(0.98;1.25) |
| In-Hospital Deaths |  |  |  |  |
| CVD | 49<br>(45;54) | 56<br>(51;60) | -6.3 | 0.89<br>(0.79;0.99) * |
| ACS | 7<br>(5;8) | 9<br>(7;11) | -2.2 | 0.76<br>(0.57;1.01) |
| Stroke | 16<br>(14;18) | 17<br>(14;19) | -0.7 | 0.96<br>(0.79;1.18) |
| HF | 13<br>(11;15) | 12<br>(10;14) | 0.5 | 1.04<br>(0.83;1.31) |
| % In-Hospital Deaths |  |  |  |  |
| CVD | 6<br>(5;6) | 5<br>(5;5) | 0.9 | 1.16<br>(1.04;1.3)* |
| ACS | 4<br>(3;4) | 4<br>(3;5) | -0.2 | 0.96<br>(0.71;1.28) |
| Stroke | 10<br>(9;12) | 11<br>(9;13) | -0.8 | 0.93<br>(0.76;1.13) |
| HF | 8<br>(6;9) | 7<br>(6;8) | 0.6 | 1.08<br>(0.86;1.36) |

ACS: acute coronary syndrome, CVD: cardiovascular disease, HF: heart failure, ICU: intensive care unit

**Table 2** shows the risk ratios and their 95% confidence intervals for total CVD in subgroup analysis. Regarding sex-stratified analysis, no differences between men and women were found, expect that the increase in proportions of ICU utilization and in-hospital mortality were only significant for women. The analysis by age groups revealed that the increase in home deaths was only significant for older adults (≥ 60y). Of note, mortality rates were found to be at least 10 times higher in the older age group, what may have precluded finding statistical significance in the younger group, for which the direction of changes were the same as for the older age group (**Supplementary Material 4**).

**Table 2:** Risk ratios for total cardiovascular disease outcomes and their 95% confidence intervals in subgroup analysis, according to sex, age group, health vulnerability, and pandemic phase, in Belo Horizonte, Brazil, from epidemiological weeks 10-48, 2020, compared to the mean of epidemiological weeks 10-48, 2015-2019.

|  | Deaths | Deaths at Home | % Deaths at Home | Hosp. Admissions | ICU | % ICU | In-Hospital Deaths | % In-Hospital Deaths |
| --- | --- | --- | --- | --- | --- | --- | --- | --- |
| **Sex** |  |  |  |  |  |  |  |  |
| Feminino | 0.98<br>(0.91;1.06) | 1.35<br>(1.17;1.56)* | 1.35<br>(1.17;1.56)* | 0.69<br>(0.66;0.72)* | 0.87<br>(0.81;0.94)* | 1.26<br>(1.17;1.35)* | 0.86<br>(0.73;1.02) | 1.2<br>(1.02;1.42)* |
| Masculino | 1.04<br>(0.97;1.11) | 1.29<br>(1.13;1.47)* | 1.27<br>(1.11;1.44)* | 0.84<br>(0.81;0.87)* | 0.88<br>(0.83;0.94)* | 1.05<br>(0.99;1.12) | 0.91<br>(0.78;1.06) | 1.14<br>(0.98;1.32) |
| **Age Groups** |  |  |  |  |  |  |  |  |
| 30-59 | 0.91<br>(0.79;1.04) | 1.09<br>(0.85;1.4) | 1.2<br>(0.93;1.54) | 0.66<br>(0.63;0.69)* | 0.86<br>(0.79;0.92)* | 1.29<br>(1.19;1.39)* | 0.8<br>(0.63;1.01) | 1.2<br>(0.95;1.53) |
| 60+ | 1.03<br>(0.97;1.09) | 1.37<br>(1.23;1.52)* | 1.33<br>(1.2;1.48)* | 0.83<br>(0.8;0.86)* | 0.89<br>(0.84;0.94)* | 1.07<br>(1.01;1.14)* | 0.91<br>(0.8;1.04) | 1.1<br>(0.97;1.25) |
| **Vulnerability** |  |  |  |  |  |  |  |  |
| Low | 0.95<br>(0.86;1.05) | 1.21<br>(1.01;1.45)* | 1.25<br>(1.04;1.49)* | 0.72<br>(0.67;0.78)* | 0.82<br>(0.73;0.92)* | 1.12<br>(0.99;1.26) | 1.02<br>(0.76;1.38) | 1.32<br>(0.98;1.79) |
| Medium | 1.01<br>(0.93;1.09) | 1.32<br>(1.13;1.53)* | 1.32<br>(1.13;1.53)* | 0.76<br>(0.73;0.79)* | 0.89<br>(0.83;0.95)* | 1.18<br>(1.1;1.26)* | 0.88<br>(0.75;1.03) | 1.2<br>(1.03;1.41)* |
| High | 1.04<br>(0.94;1.14) | 1.45<br>(1.21;1.74)* | 1.41<br>(1.17;1.69)* | 0.81<br>(0.78;0.85)* | 0.9<br>(0.84;0.97)* | 1.13<br>(1.05;1.21)* | 0.82<br>(0.69;0.99)* | 1.13<br>(0.95;1.36) |
| **Pandemic Phase** |  |  |  |  |  |  |  |  |
| 1° Period | 1.01<br>(0.92;1.12) | 1.37<br>(1.15;1.63)* | 1.35<br>(1.13;1.6)* | 0.77<br>(0.73;0.81)* | 1.03<br>(0.95;1.12) | 1.34<br>(1.24;1.45)* | 0.87<br>(0.7;1.06) | 1.12<br>(0.91;1.38) |
| 2° Period | 0.96<br>(0.86;1.06) | 1.35<br>(1.11;1.63)* | 1.4<br>(1.16;1.7)* | 0.75<br>(0.71;0.8)* | 0.8<br>(0.73;0.89)* | 1.07<br>(0.97;1.18) | 0.81<br>(0.64;1.02) | 1.07<br>(0.85;1.36) |
| 3° Period | 1.03<br>(0.95;1.12) | 1.28<br>(1.1;1.48)* | 1.24<br>(1.07;1.44)* | 0.76<br>(0.73;0.8)* | 0.82<br>(0.76;0.87)* | 1.07<br>(1;1.14) | 0.95<br>(0.8;1.11) | 1.24<br>(1.05;1.46)* |

ICU: intensive care unit

In the analysis according to social vulnerability groups, the rise in deaths at home during the 2020 pandemic period occurred in the three HVI groups, but with an increasing gradient from the low to medium, and high vulnerability groups, as also depicted in **Figure 1**.

**Figure 1:**
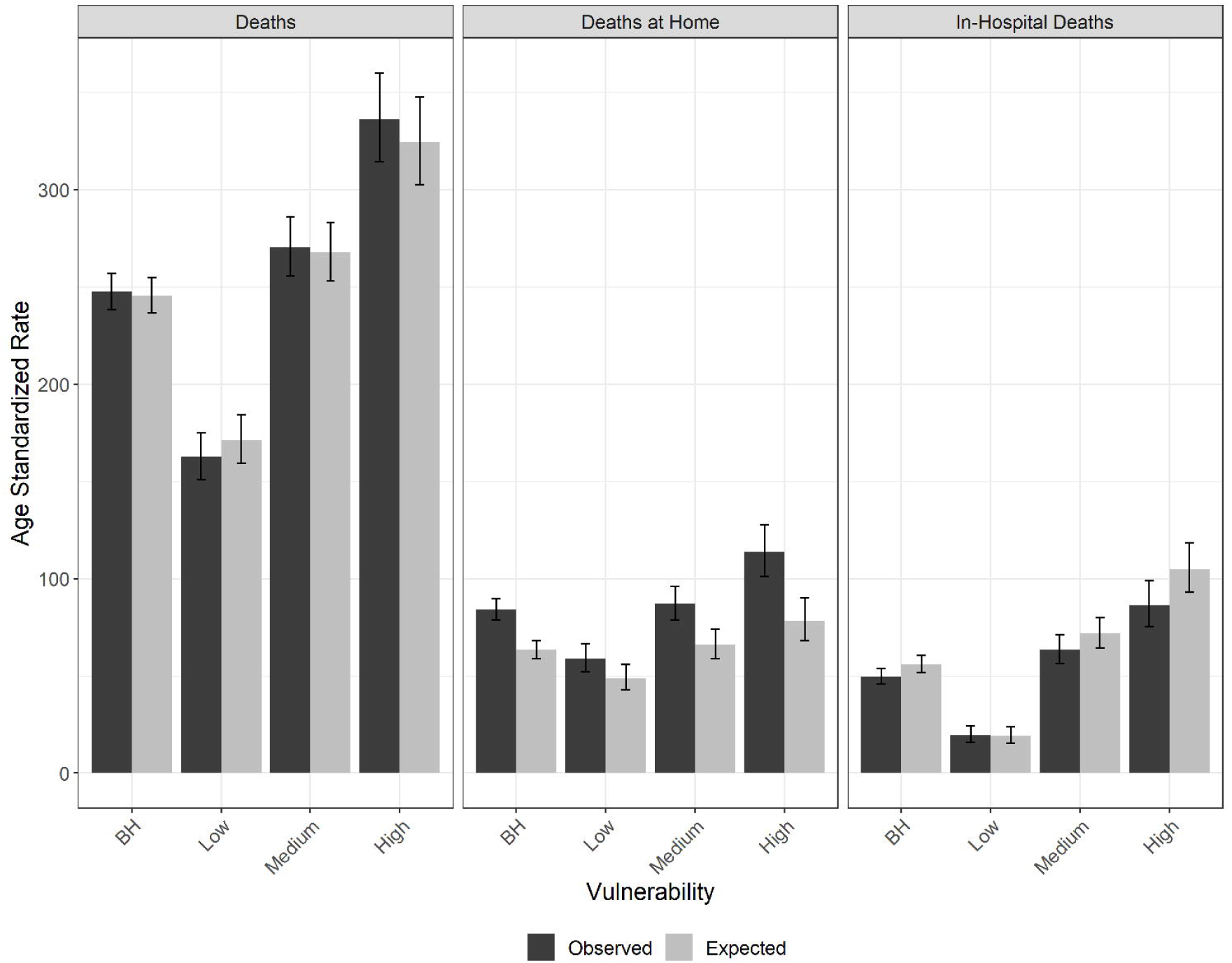
Age-standardized death, death at home, and in-hospital death rates per 100,000 inhabitants, for cardiovascular diseases, observed in 2020 and expected (mean of 2015-2019) for epidemiological weeks 10-48, with 95% confidence intervals. Belo Horizonte, MG, Brazil

Lastly, **Figure 2** and **Table 2** reveals that there was in increase in deaths at home in all pandemic phases during the analysed period. However, the profile of hospitalizations suggest different drivers for the excess mortality at home: while there was a decrease for hospitalization in all phases, ICU admission rates only decreased in the second and third phases, possibly due to competing demands. In fact, the proportion of ICU utilization increased in the first period of the pandemic.

**Figure 2:**
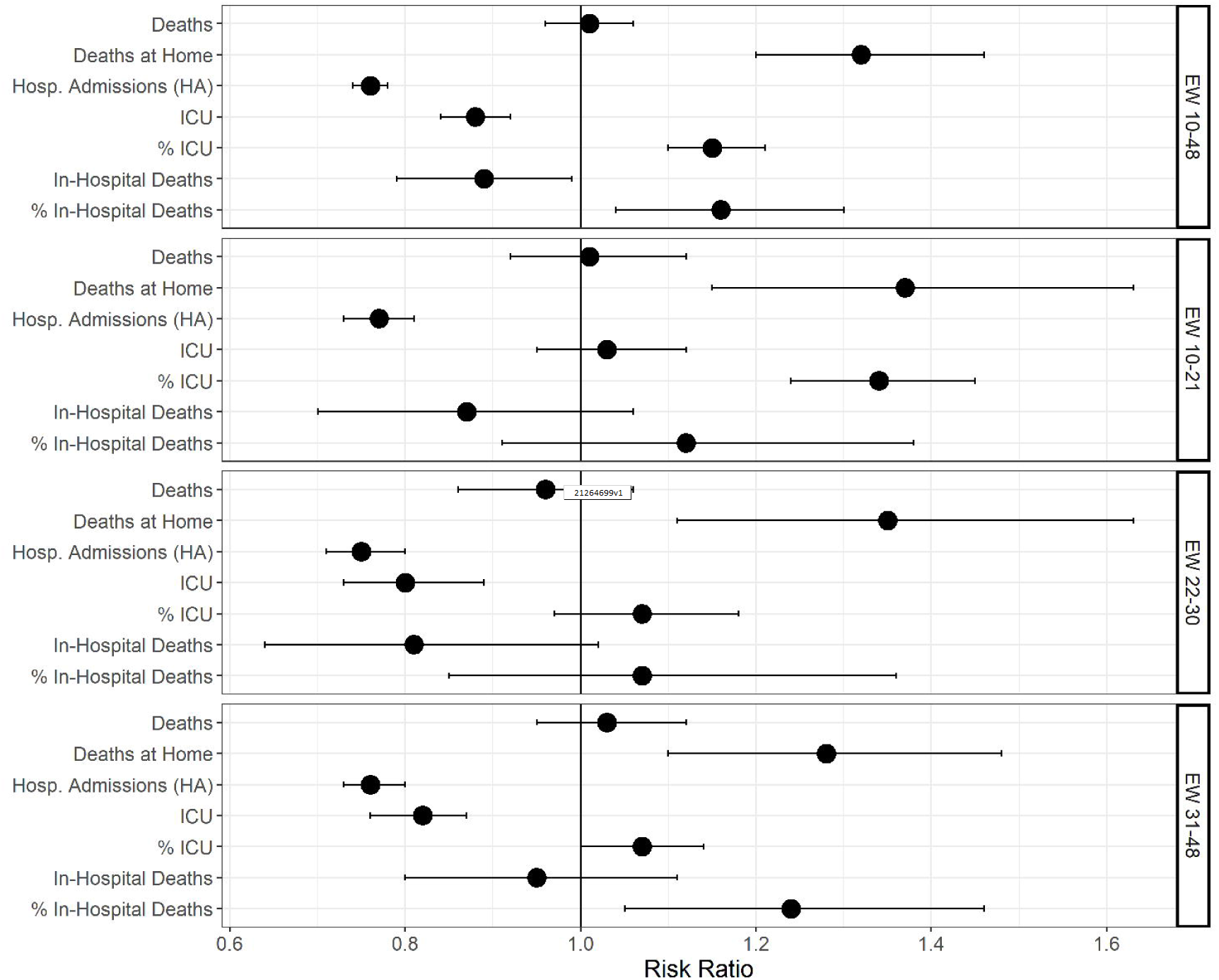
Risk ratios expected (mean of 2015-2019) and observed in 2020 for age-standardized rates per 100,000 inhabitants or proportions for cardiovascular disease outcomes for epidemiological weeks 10-48. Belo Horizonte, MG, Brazil

## Discussion

During the 2020 pandemic period, CVD deaths did not increase in Belo Horizonte, although there was a displacement of deaths, which occurred more at homes than at hospitals, even though the proportion of in-hospital deaths increased. These results suggest that there was a deferral of CVD care in hospitals, with higher in-hospital lethality, possibly due to greater clinical severity of hospitalized patients, or less adherence to evidence–based treatments because of disrupted pathways of care and overwhelmed hospitals. Importantly, this unfavorable impact on CVD care in the city was differential, mainly affecting older adults, and with an increasing trend in the more socially vulnerable groups, widening health gaps. Moreover, the changes in the location of deaths occurred during the whole 2020 pandemic period, although hospitalization and ICU utilization patterns suggest that these changes may have been led by different drivers: avoidance of hospitals at the first phase, and additional competition for beds in the second and third phases. Taken together, our study brings essential information for preparing the local health system for future waves of this or different pandemics, and to plan actions to mitigate the rebound effect resulting from the disrupted CVD care in the city.

Previous studies have shown an indirect effect of the COVID-19 pandemic in CVD care. First, studies revealed a reduction in hospitalizations, procedures, and/or consultations for CVD in high,^26–30^ and low-and middle income countries (LMIC),^31,32^ then an increase in out-of-hospital deaths.^33,34^ Moreover, a rise in the proportion of in-hospital mortality for CVD was reported, being this effect larger where greater decline in admission rates occurred, suggesting hospitalizations of sicker patients or delivery of worse quality of care, corroborating our findings.^35,36^ In parallel, other studies revealed that excess mortality in the pandemic period could not be explained exclusively by COVID-19 deaths, and excess CVD deaths was found in some countries, including Brazil, which is not surprising, knowing the beneficial effect of in-hospital timely treatment for the most lethal cardiovascular conditions, such as ACS.^37–39^

However, even though the effect of the pandemics in CVD deaths had already been investigated in six Brazilian capital cities, the findings could not be extended to Belo Horizonte due to the heterogeneity of the pandemics in Brazil. The cited study revealed that the cardiovascular mortality increase during the pandemics was of greater magnitude in the less developed cities, where a health system collapse occurred.^7^ Moreover, the effect of mitigation policies or the differential impact according to individual-level factors, both fundamental to guide health policies, needed to be investigated.

In our analysis in Belo Horizonte, CVD death rates did not increase in the period. This finding could be attributed to Belo Horizonte having a smaller incidence of COVID-19 - and as a result, a lower overall impact of the pandemics compared to other Brazilian cities - or a better organization of the health system to cope with competing demands.^11^ In fact, previous studies in the US and Brazil, showed that deaths from CVD increased more during the pandemics in areas of higher incidence of COVID-19.^2,7^ In addition, the health system collapse described in other Brazilian cities, such as Manaus, did not occur in Belo Horizonte in 2020.^11^ However, when we comprehensively analyze other cardiovascular outcomes in Belo Horizonte, we clearly found a disruption of CVD care in the city, with a rise in deaths at home – also seen in other countries-,^5,40^ and significant declines in hospitalizations and in-hospital deaths, even though there was a rise in lethality. Looking at this overall pattern of CVD outcomes during the period in Belo Horizonte, an alternative for not finding an increase in CVD death rates may be lack of power to detect differences during the studied period. Importantly, individuals living with CVD are at increased risk of death due to COVID-19, and thus there may be an effect of competitive causes for this group in the pandemics period between COVID-19 and CVD deaths.^41^

To further understand the effect on CVD care, we stratified the analysis by the main causes of cardiovascular death or hospitalization in the country, and interestingly we found more adverse effect on the mortality by HF, compared to ACS and stroke. Of note, not only deaths at home increased for HF in greater magnitude, but also the overall HF death rate rose. This finding contrasts with data from the US, where excess deaths for ischemic and hypertensive diseases were responsible for CVD excess mortality, and no increase in deaths by heart failure was observed, leading the authors to describe a potential bigger unfavorable effect on acute cardiovascular conditions.^2^ However, this may differ in Brazil, where the care of chronic CVD may have been more adversely affected. During the early 2020 pandemic period, although the PHC centers designed protocols for attending the patient with CVD,^42^ any contact with the health system was, in fact, discouraged by national authorities, particularly for older individuals, what may have resulted in the decompensation of individuals living with HF.^43^ A survey conducted in April and May, 2020, with 45,000 Brazilian adults, revealed a decrease in health services’ demand, and a reported difficulty in scheduling exams, consultations, elective hospitalization, and procedures, reflecting the “stay at home” order from the first pandemics’ phase, as well as the fear of contracting COVID-19.^44^ Indeed, our study showed that hospitalizations for HF did not decline in the period, suggesting that HF decompensation were still highly demanding hospitals. Alternatively, HF may have been assigned as cause of death in the absence of a more specific diagnosis, harder to make at home or early after a hospital admission due to the lack of diagnostic tools.

Importantly, our study corroborates previous data from Brazil and other countries, which reveal that the pandemic exacerbated health inequities.^6,9,45,46^ More excess mortality and COVID-19 confirmed deaths have been reported in the most vulnerable groups, according to race or neighbourhood.^4,6,9^ Herein, we additionally describe that the adverse impact of the pandemic on CVD was also greater in the most vulnerable individuals: deaths at home presented an increasing trend from groups of low to high health vulnerability. This finding depicts the inequality chain in which the most vulnerable individuals live, with higher prevalence of unhealthy behaviours and cardiovascular risk factors, such as obesity and hypertension, which lead to higher prevalence of CVD.^47,48^ Besides that, although Brazil has a universal healthcare system and the quality of PHC in Belo Horizonte has been rated as one of the best in the countries’ capital cities,^11^ PHC was not promptly adapted to deliver care for CVD in the pandemic context, as already described. Moreover, gaps in accessibility exist, such as having more public hospitals located in the less vulnerable areas of the city - a disadvantage for the treatment of time-dependent conditions, such as ACS, for those living in more vulnerable areas.^39^ Additionally, other barriers to CVD care for more socially vulnerable groups may include impaired access to information, and to alternative pathways of care, such as telemedicine.^49^ From a macro-level perspective, lower education, health literacy, and economic constraints may have also been determinants of lower access to health care to the most vulnerable, reinforcing disparities.^50^

Our study has limitations. Because COVID-19 tests were not widely available in the community setting, out-of-hospital deaths may have been misdiagnosed, either over or underestimated deaths by CVD. Moreover, quality of death certificates may have worsen during the pandemic.^51^ However, no changes in the pattern of deaths under ICD-10 Chapter XVIII, which includes ill-defined causes of deaths, was found in our analysis, suggesting that this was not a major issue. Importantly, because our aim was to understand the indirect impact of the pandemic on CVD, we did not include in the present analysis the cardiovascular deaths related to COVID-19. As such, if COVID-19 was considered the underlying cause of death according to the World Health Organization protocols, and there was an associated cardiovascular outcome in the death certificate, this outcome was not included in the present analysis,^22,52^ as it would require an analysis of multiple causes of death.^52^ Another limitation refers to the fact that only public hospitalizations were analysed, due to availability of data. However, there was no substantial changes in the number of individuals with private health insurance in the city during the period.^16^ While the definition of out-of-hospital cause of death was described as a potential limitation, the same does not occur for in-hospital mortality, as hospitalizations for COVID-19 had to be confirmed by laboratory tests (RT-PCR or serology) that, although not widely available at the community level, were available for in-hospital diagnosis in Belo Horizonte.^53,54^

The strengths of our analysis include a thorough analysis of data quality that supported our decision to only include information until EW 48, to minimize the effect of reporting delays. Moreover, the age standardization of data was fundamental to reveal the difference among the HVI groups, since the higher the HVI, the greater is the proportion of younger individuals, masking the larger increase in mortality in the most vulnerable groups, if crude mortality rates were used.^9^ Lastly, the comprehensive analysis of the pandemic’s impact on CVD with a focus in a city allowed us to understand the effect of the pandemic course, including that of mitigation policies.

Taken together, our analysis brings important messages for health professionals and stakeholders: tools to preclude disruption of CVD care must be developed and implemented during a health crisis. Alternative pathways of care, such as homecare, active search, telemedicine, digital health tools, or reorganization of PHC centres to partly devote them to chronic diseases management are options to keep track of chronic patients and stimulate the continuation of treatment.^49,50,55^ Enhancement of educational campaigns to increase recognition of acute CVD symptoms, highlighting the importance of seeking emergency medical services in their presence are another essential strategy.^5^ Additionally, health promotion should also be emphasized to counteract the increase of unhealthy behaviours that occurred during the pandemics, if we intend to mitigate the deleterious effect on cardiovascular health in the years to come. A Brazilian national survey found an increase in the prevalence of sedentary behaviour, smoking, consumption of alcohol, and of a diet rich in ultra-processed foods.^56–58^ From the societal perspective, investments in social protection policies and education are essential to break the chain of inequalities.

## Conclusion

During the 2020 COVID-19 pandemic period, CVD care was disrupted in Belo Horizonte city. CVD deaths were displaced to homes instead of hospitals during the period, concomitantly with a decrease in hospitalizations, suggesting that there was a deferral of hospital care for CVD. This unfavourable impact of COVID-19 pandemic was greater for older adults, and the most socially vulnerable group, exacerbating health inequalities in the city.

## Data Availability

All data produced in the present work are contained in the manuscript

## Authors’ contributions

Brant, LC designed the work; supervised the data analysis and interpretation, and drafted the first version of the manuscript; Passos VMA designed the work; supervised the data analysis and interpretation, and critically revised the manuscript; Machado, IE; Correa PR; Santos MR; Malta DC; Ribeiro, ALP designed the work and collaborated in and data acquisition and interpretation, and critically revised the manuscript; Pinheiro, PC designed the work and did the statistical analysis, Souza MFM collaborated in the design of the work, in the interpretation of data, and critically revised the manuscript; and all authors approved this version of the manuscript.

