## Supplementary material for "Cardiovascular Mortality during the COVID-19 Pandemics in a Large Brazilian City: a Comprehensive Analysis": Sup_Material_1

|  |  | Processing Month | | | | | | | | | | | | | | | | |
| --- | --- | --- | --- | --- | --- | --- | --- | --- | --- | --- | --- | --- | --- | --- | --- | --- | --- | --- |
|  |  | 2019-01 | 2019-02 | 2019-03 | 2019-04 | 2019-05 | 2019-06 | 2019-07 | 2019-08 | 2019-09 | 2019-10 | 2019-11 | 2019-12 | 2020-01 | 2020-02 | 2020-03 | 2020-04 | 2020-05 |
| Hospital Admission Month | 2019-01 | 32.32 | 41.01 | 16.58 | 8.25 | 1.24 | 0.36 | 0.06 | 0.02 | 0.02 | 0.02 | 0.02 | 0.01 | 0.01 | 0.01 | 0.01 | 0.01 | 0.01 |
|  | 2019-02 | 0 | 33.1 | 43.07 | 14.85 | 7.32 | 1.33 | 0.29 | 0.03 | 0 | 0.01 | 0 | 0 | 0 | 0 | 0 | 0 | 0 |
|  | 2019-03 | 0 | 0 | 33.81 | 40.45 | 16.94 | 7.15 | 1.23 | 0.32 | 0.07 | 0.02 | 0.02 | 0 | 0 | 0 | 0 | 0 | 0 |
|  | 2019-04 | 0 | 0 | 0 | 37.23 | 37.78 | 16.01 | 7.54 | 1.01 | 0.32 | 0.05 | 0.01 | 0.01 | 0.01 | 0.01 | 0 | 0.01 | 0.01 |
|  | 2019-05 | 0 | 0 | 0 | 0 | 35.97 | 38.93 | 16.02 | 7.43 | 1.19 | 0.3 | 0.04 | 0.01 | 0.01 | 0.01 | 0.01 | 0.01 | 0.01 |
|  | 2019-06 | 0 | 0 | 0 | 0 | 0 | 32.36 | 45.87 | 13.17 | 6.98 | 1.25 | 0.34 | 0.03 | 0 | 0 | 0 | 0 | 0 |
|  | 2019-07 | 0 | 0 | 0 | 0 | 0 | 0 | 37.18 | 42.31 | 12.08 | 6.87 | 1.07 | 0.39 | 0.09 | 0.02 | 0.01 | 0 | 0 |
|  | 2019-08 | 0 | 0 | 0 | 0 | 0 | 0 | 0 | 37.62 | 41.18 | 12.54 | 6.46 | 1.58 | 0.44 | 0.07 | 0.03 | 0.02 | 0.02 |
|  | 2019-09 | 0 | 0 | 0 | 0 | 0 | 0 | 0 | 0 | 35.65 | 44.73 | 9.96 | 7.8 | 1.39 | 0.39 | 0.06 | 0.01 | 0 |
|  | 2019-10 | 0 | 0 | 0 | 0 | 0 | 0 | 0 | 0 | 0 | 39.51 | 36.19 | 12.6 | 9.99 | 1.13 | 0.49 | 0.06 | 0.01 |
|  | 2019-11 | 0 | 0 | 0 | 0 | 0 | 0 | 0 | 0 | 0 | 0 | 37.76 | 34.52 | 17.56 | 8.39 | 1.33 | 0.37 | 0.06 |
|  | 2019-12 | 0 | 0 | 0 | 0 | 0 | 0 | 0 | 0 | 0 | 0 | 0 | 36.4 | 34.17 | 20.21 | 7.75 | 1.09 | 0.28 |

**Supplemental Material 1: Percent of hospital admissions processed in each month from the total of admissions in the respective month in 2019 and 2020, Hospital Information System, Belo Horizonte, MG, Brazil.**
