## Supplementary material for "Cardiovascular Mortality during the COVID-19 Pandemics in a Large Brazilian City: a Comprehensive Analysis": Sup_Material_2

**SUPPLEMENTAL MATERIAL 2**

**Tutorial to access data**

The following tutorial intends to help any user to access datasets from the Brazilian Ministry of Health Mortality and Hospitalization systems. It is important to emphasize that the dataset that we used in our research is part of the same information system, but it has sensitive information that unable us to share it as it is.

**HOSPITAL INFORMATION SYSTEM (SISTEMA DE INFORMAÇÕES HOSPITALARES, SIH)**

1. Access datasus webpage where the data is stored.

https://datasus.saude.gov.br/transferencia-de-arquivos/

2. In the box named ‘Download de arquivos’, select the option

‘SIHSUS – Sistema de Informações Hospitalares’

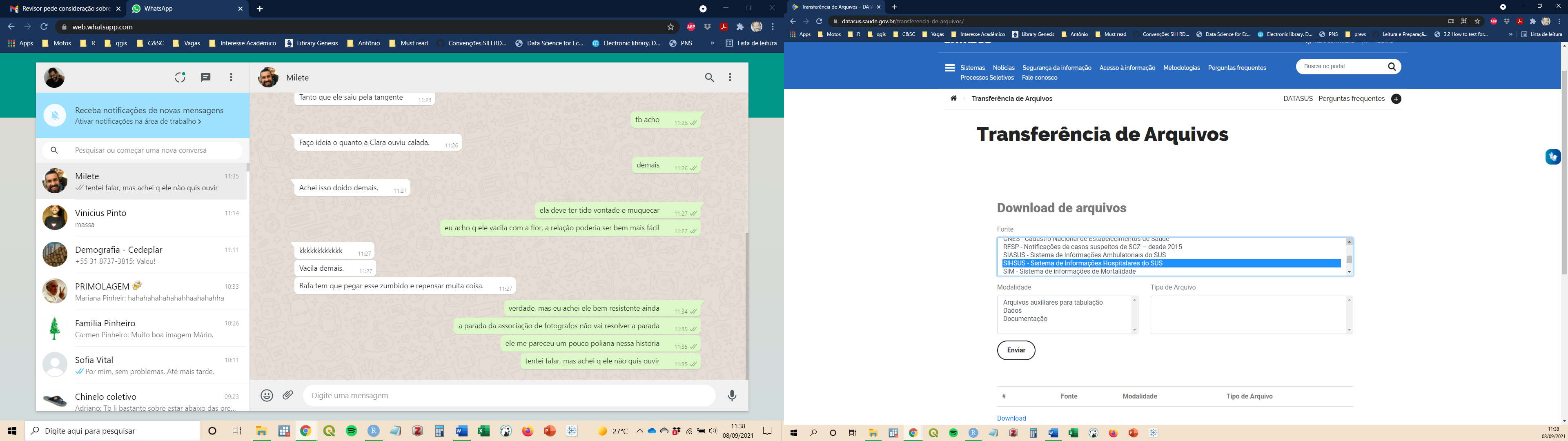

3. In the box named ‘Modalidade’, select the option ‘Dados’

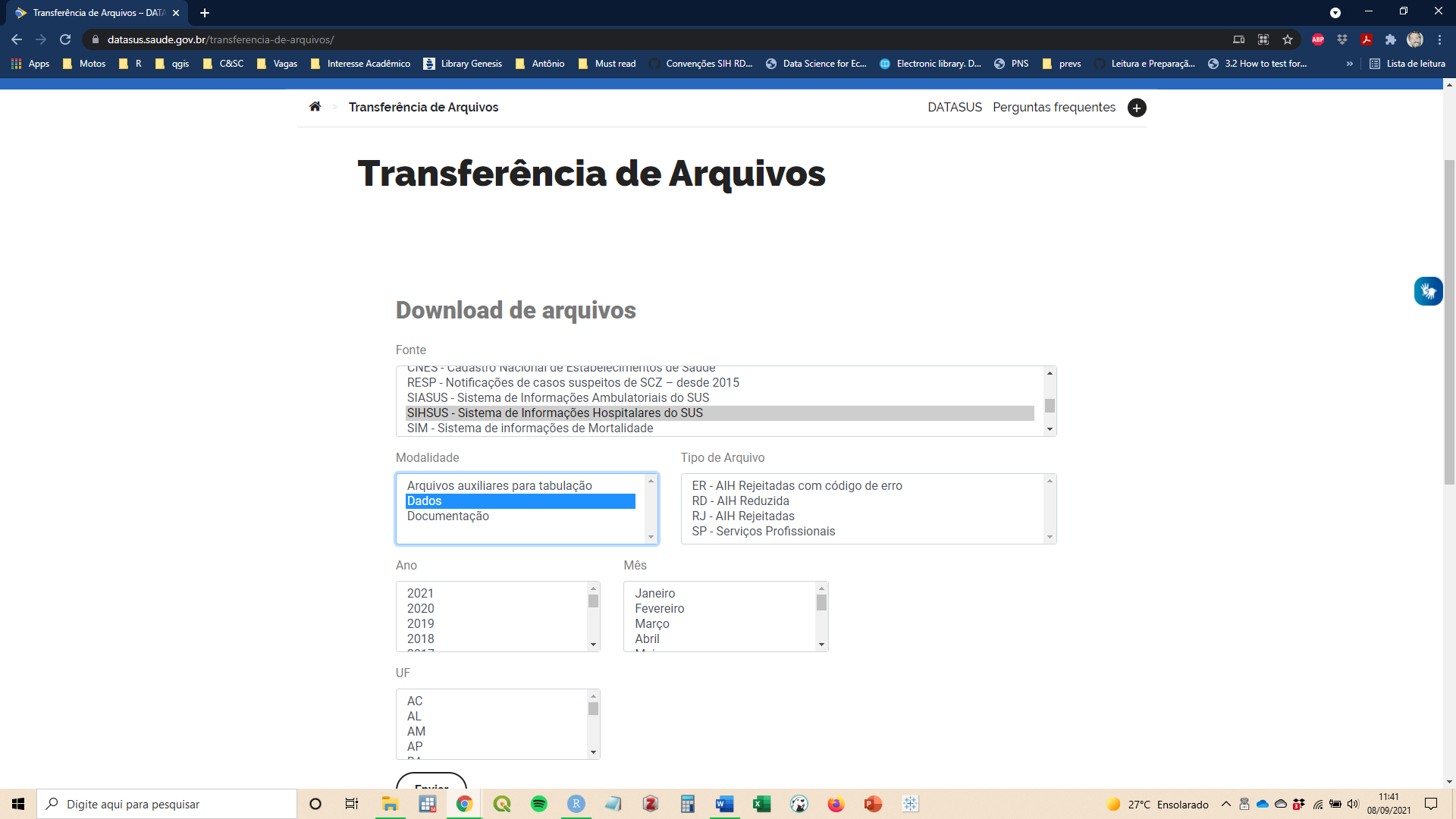

4. In the box named ‘Tipo de Arquivo’, select the option ‘RD – AIH Reduzida’

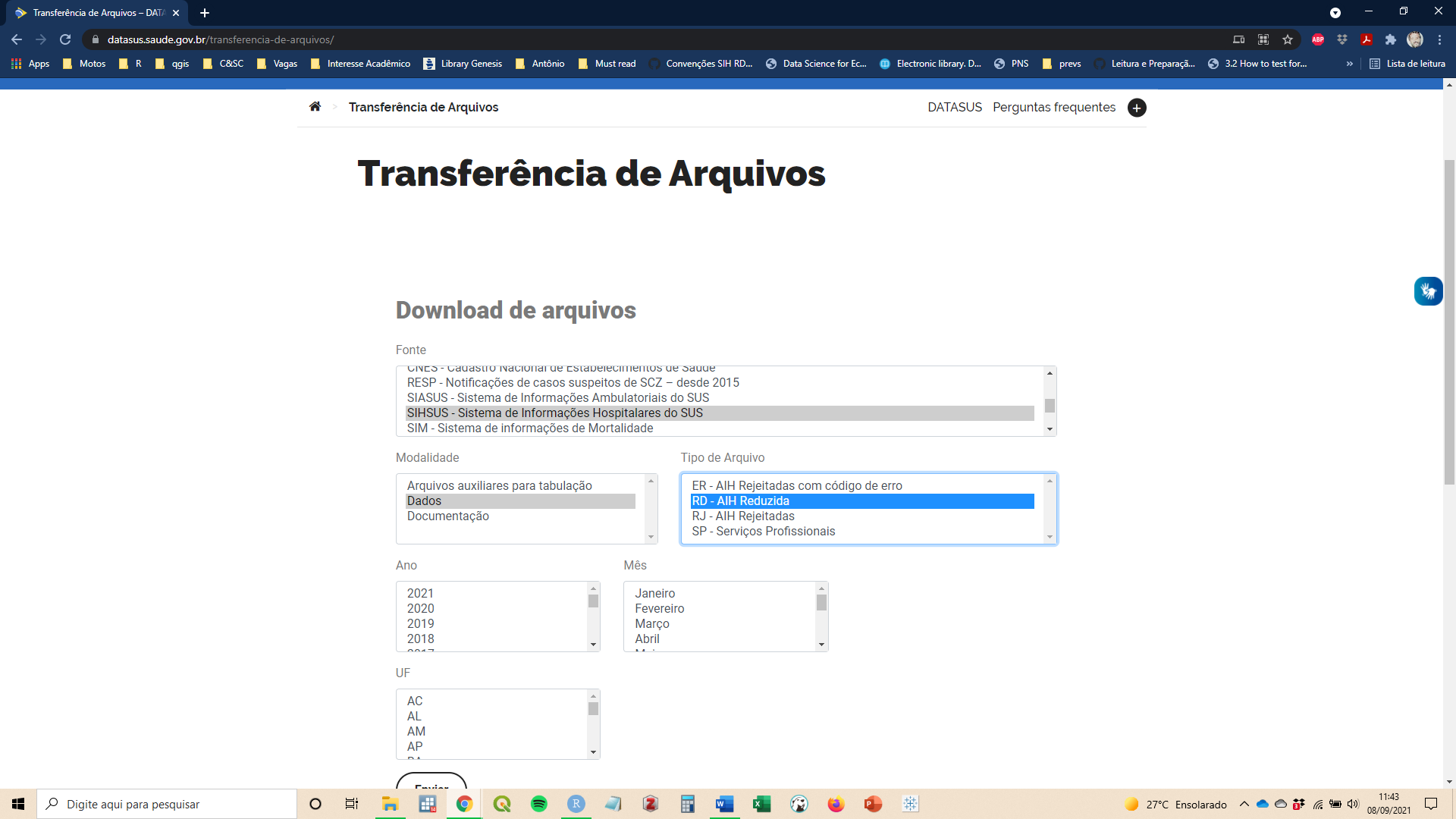

5. In the box named ‘Ano’, select the year or set of years of interest.

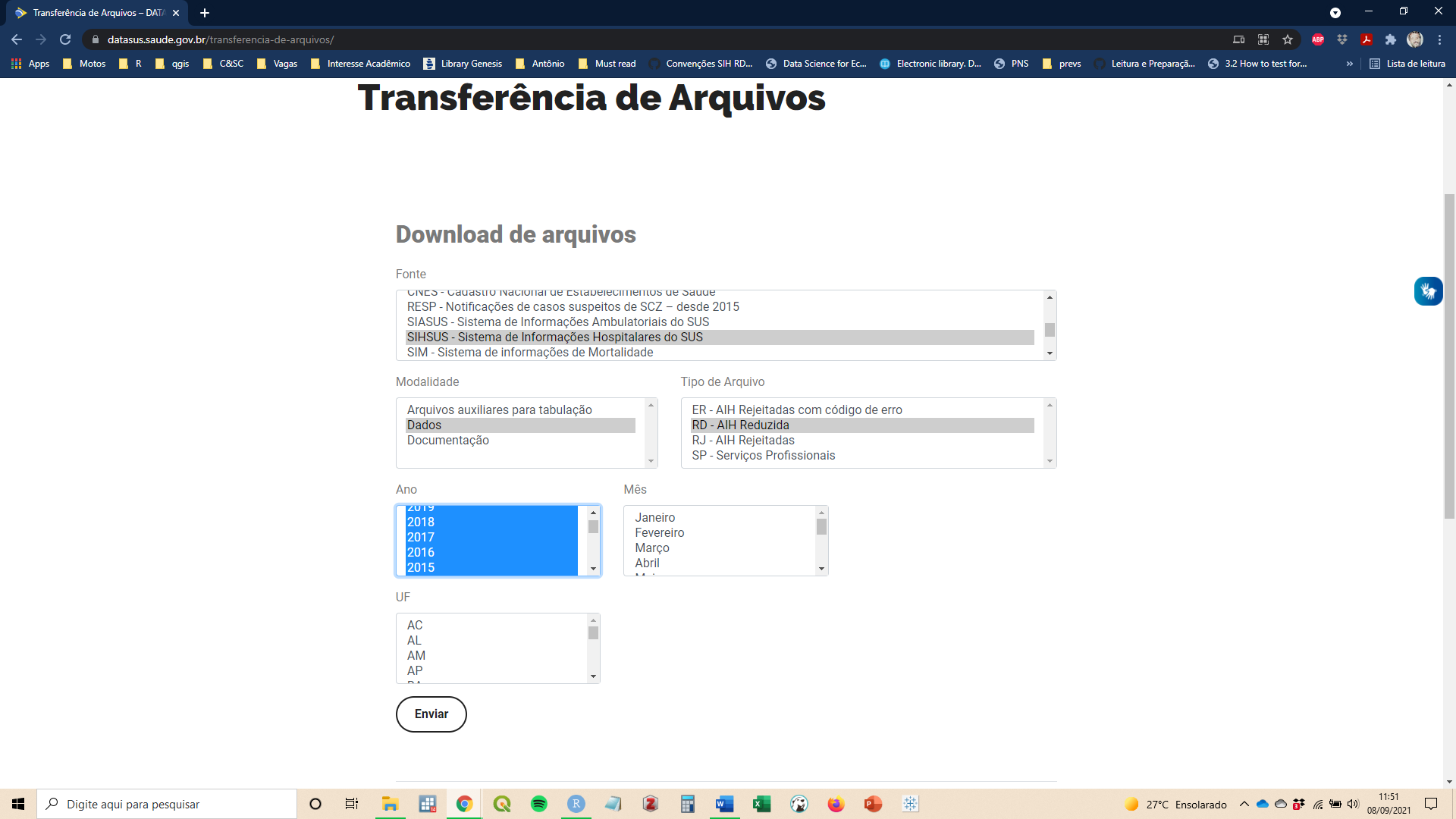

6. In the box named ‘Mês’, select the month or set of months of interest.

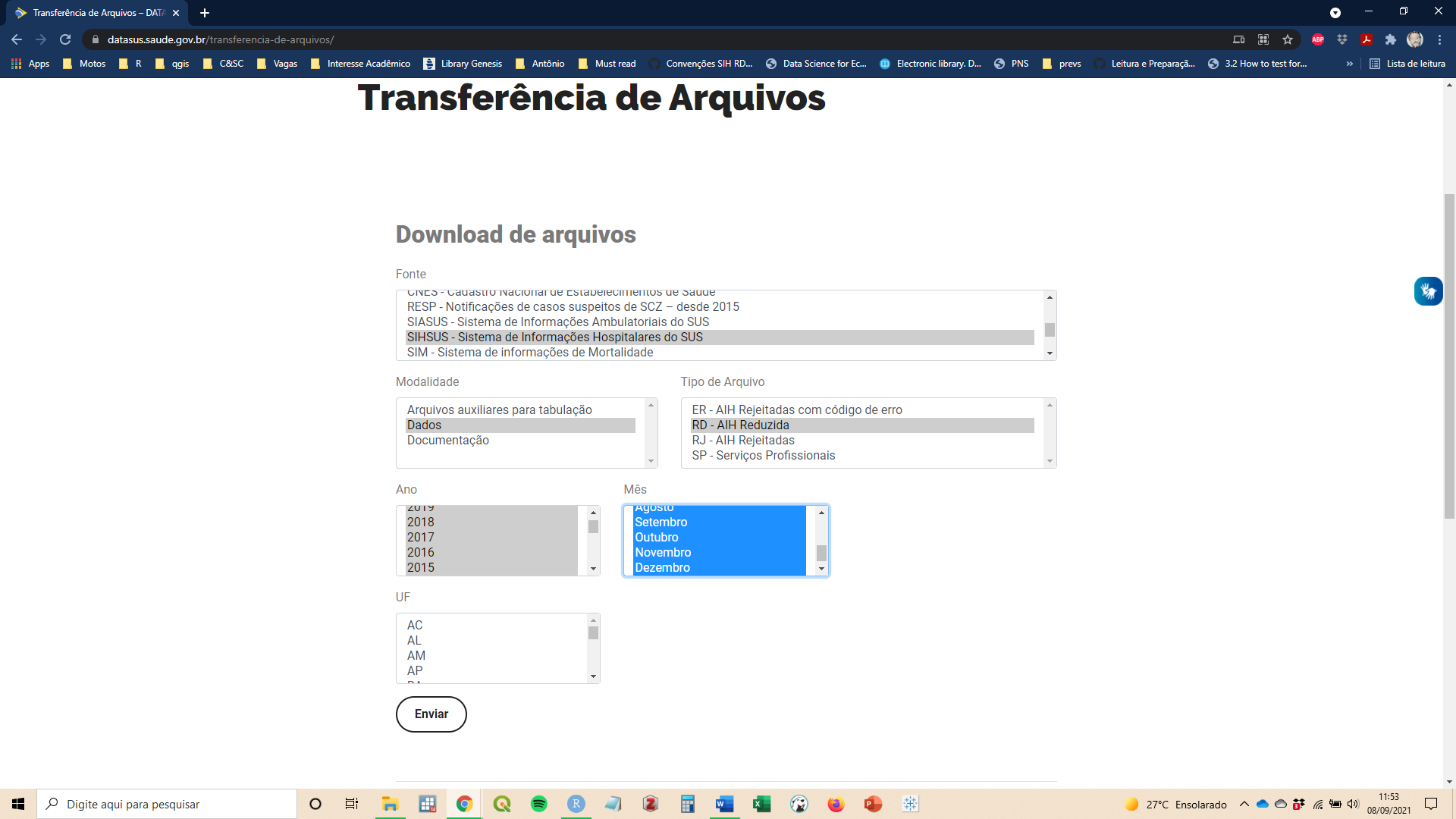

7. In the last box, named UF, choose the Brazilian state of interest. Belo Horizonte is located in Minas Gerais State, ‘MG’. Then, select ‘Enviar’.

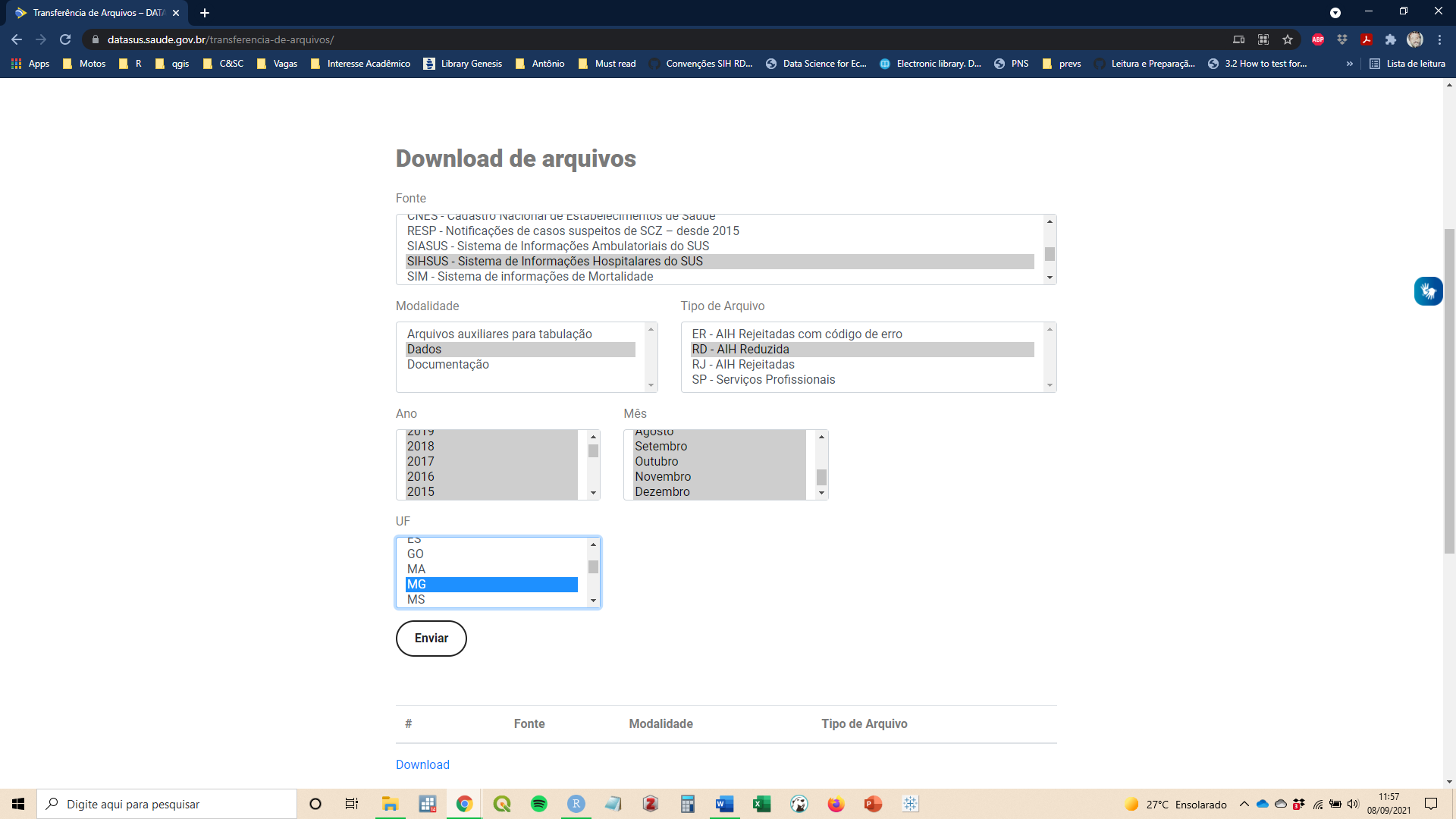

8. The selected files will be listed, at the bottom of the list you will see the download button. Files are named as ‘RD’+name of the STATE +year(YY)+month(MM)+’.dbc’

**MORTALITY INFORMATION SYSTEM (SISTEMA DE INFORMAÇÕES SOBRE MORTALIDADE, SIM)**

1. Access datasus webpage where the data is stored (same as above).

<https://datasus.saude.gov.br/transferencia-de-arquivos/>

2. In the box named ‘Download de arquivos’, select the option

‘’SIM- Sistema de Informações de Mortalidade”

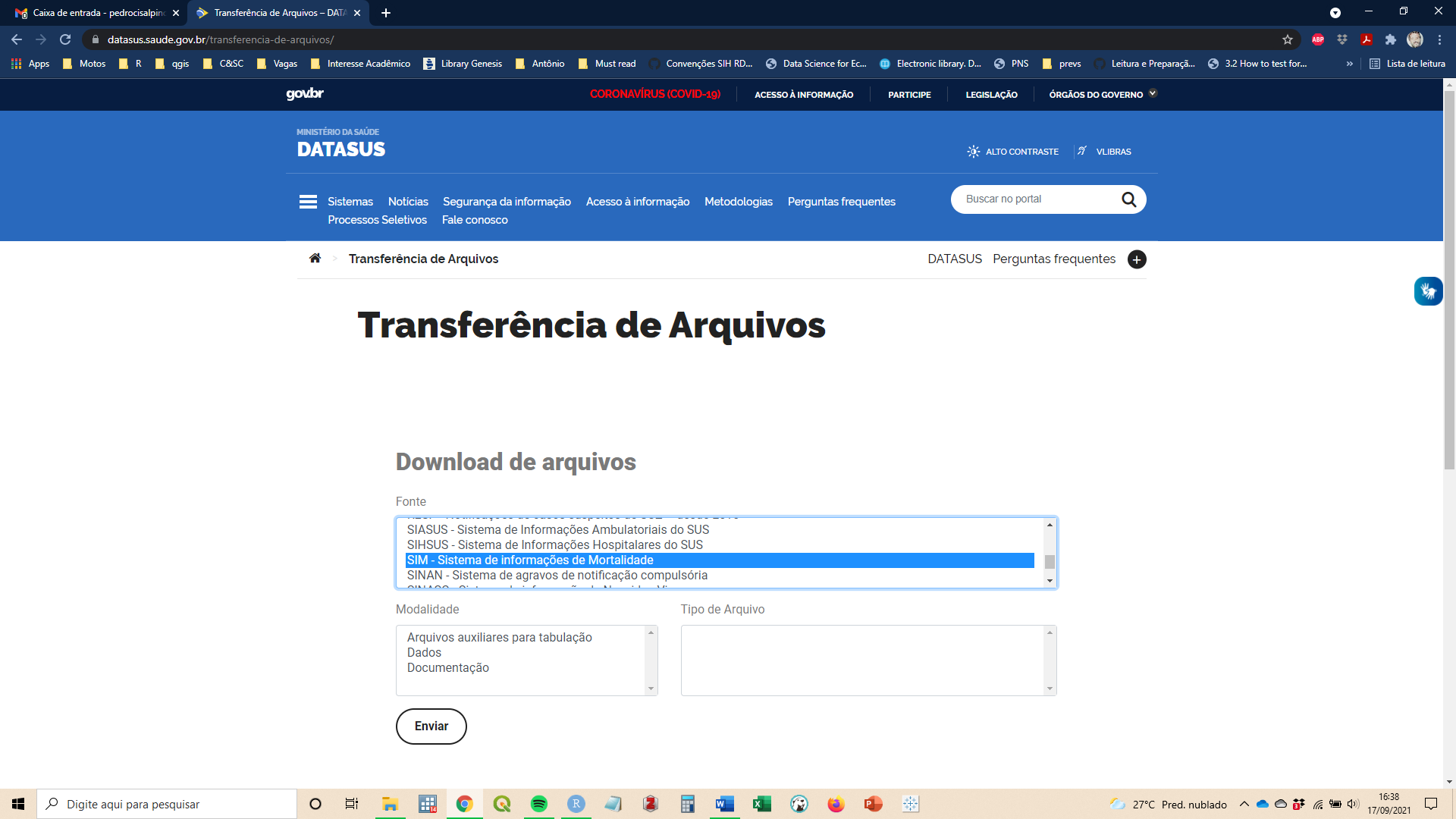

3. In the box named ‘Modalidade’, select the option ‘Dados’

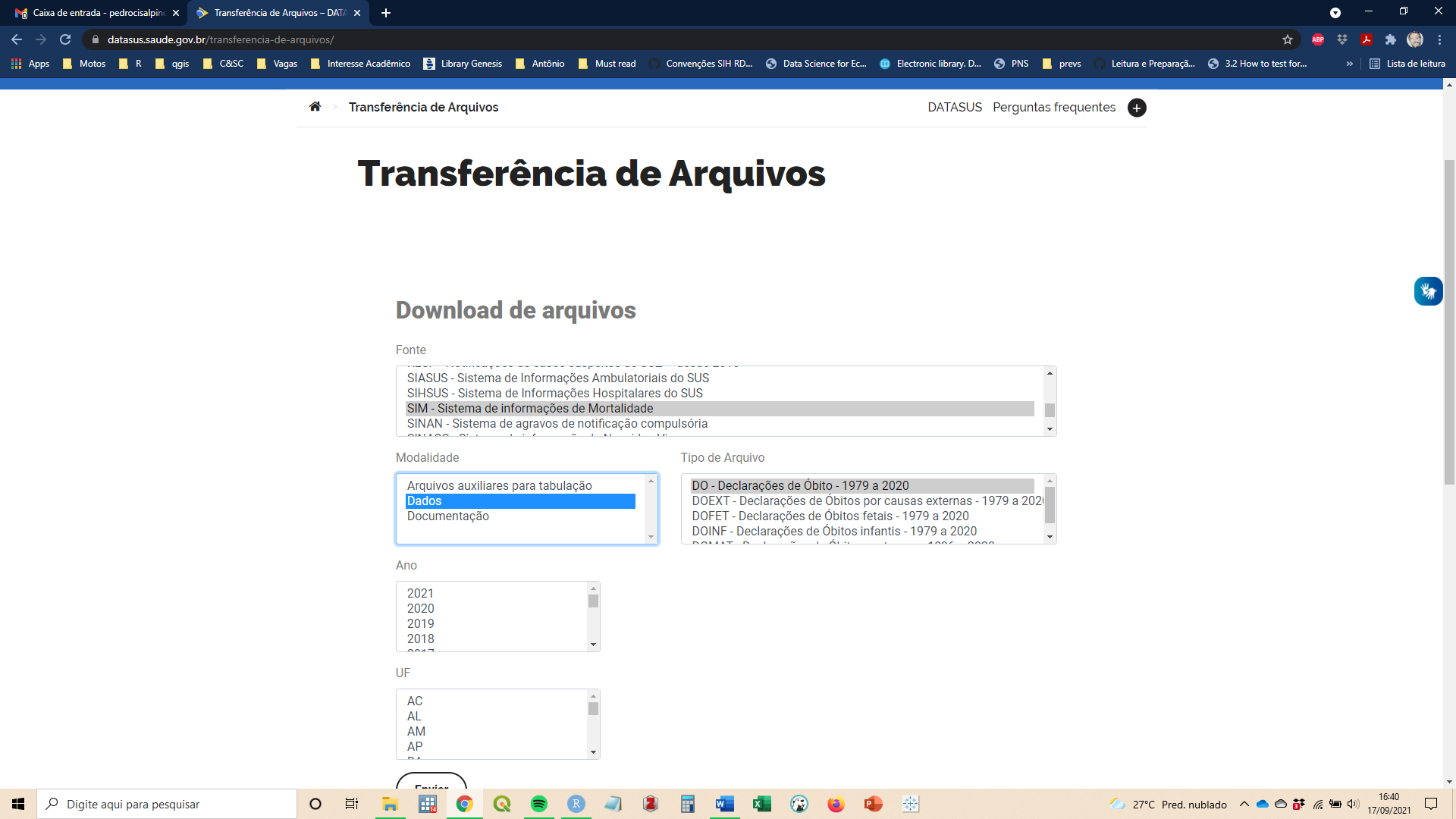

4. In the box named ‘Tipo de Arquivo’, select the option ‘DO – Declaração de Óbito – 1979 a 2020’

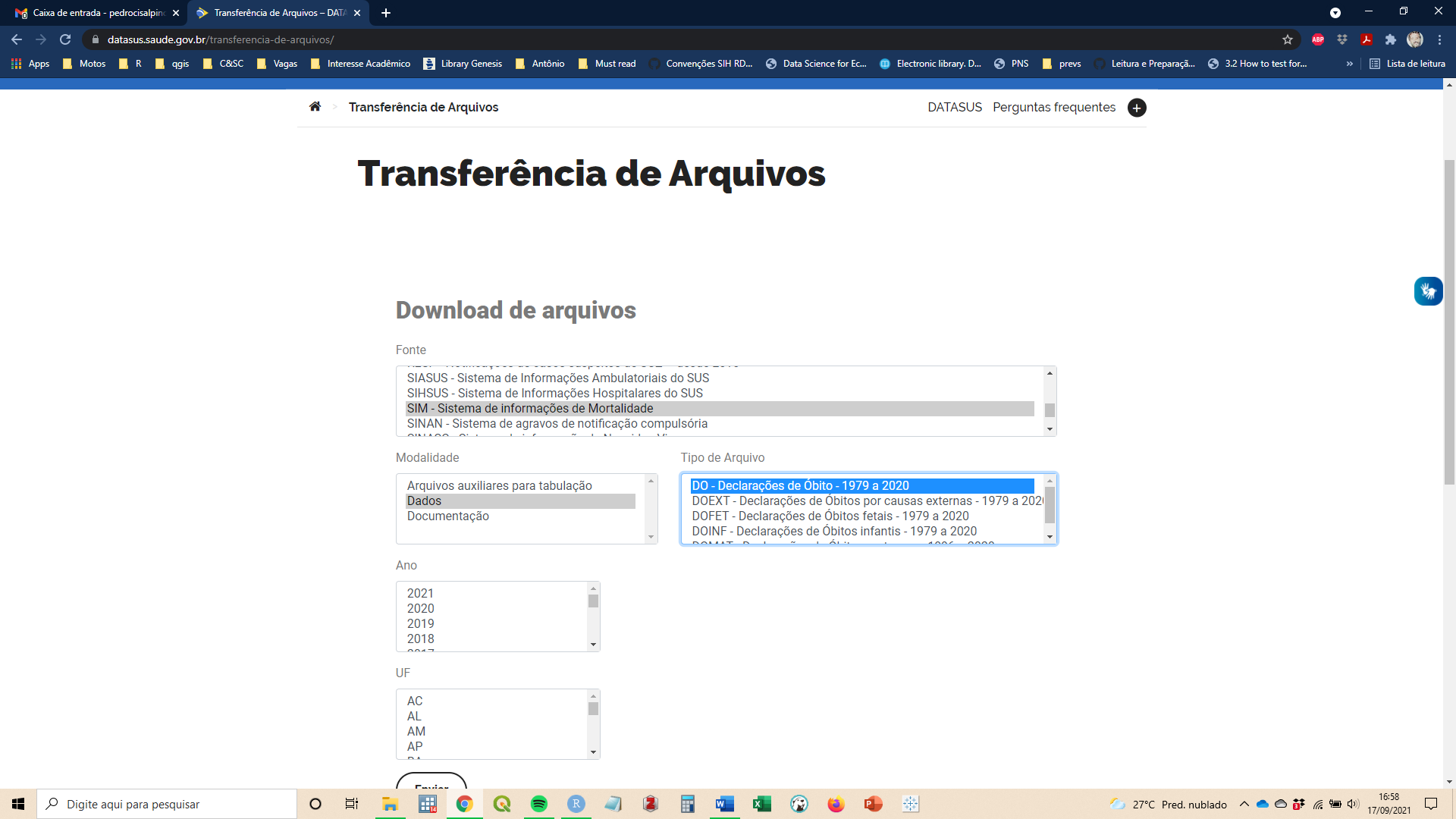

5. In the box named ‘Ano’, select the year or set of years of interest.

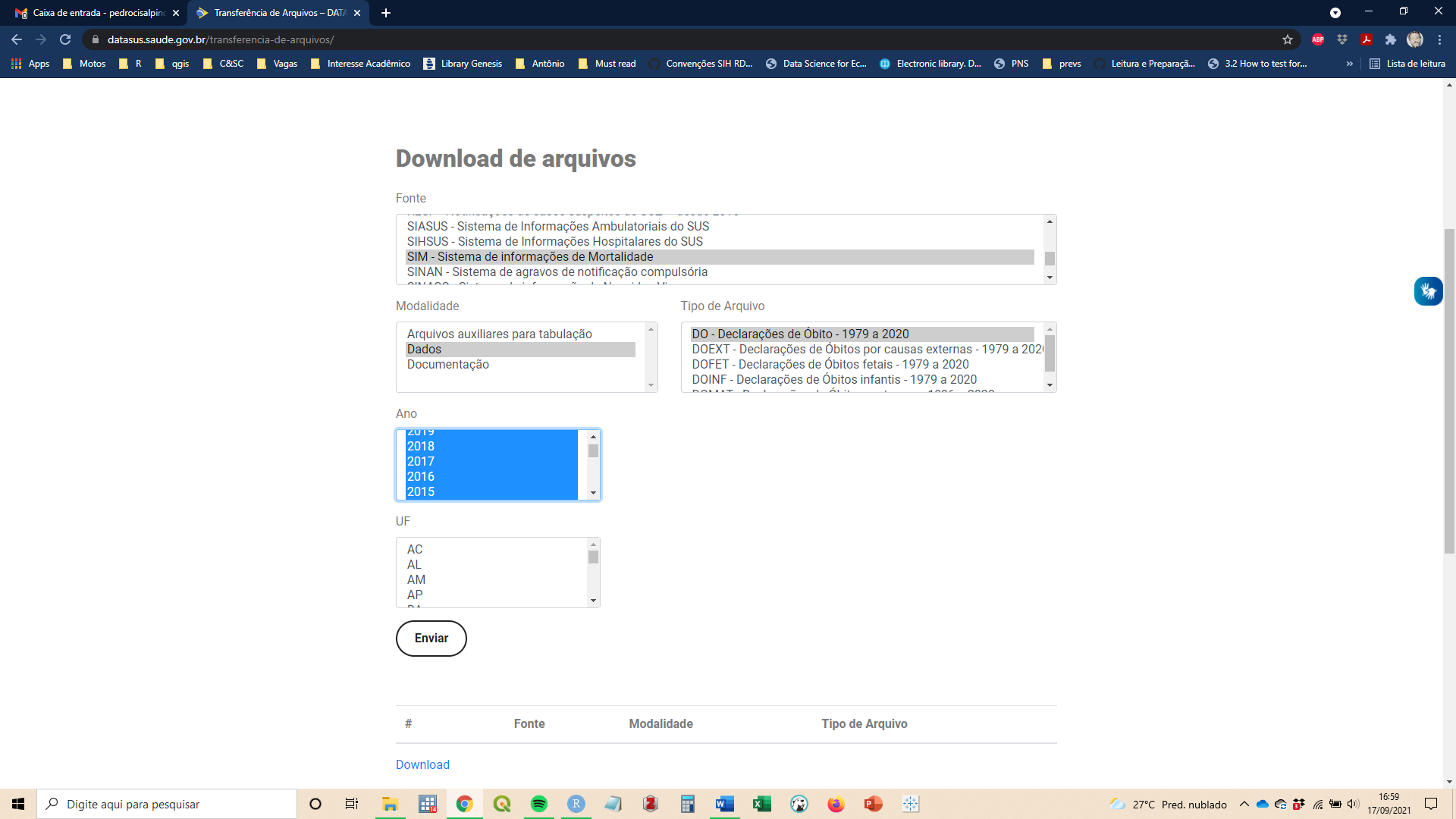

6. In the last box, named UF, choose the Brazilian state of interest. Belo Horizonte is located in Minas Gerais State, ‘MG’. Then, select ‘Enviar’.

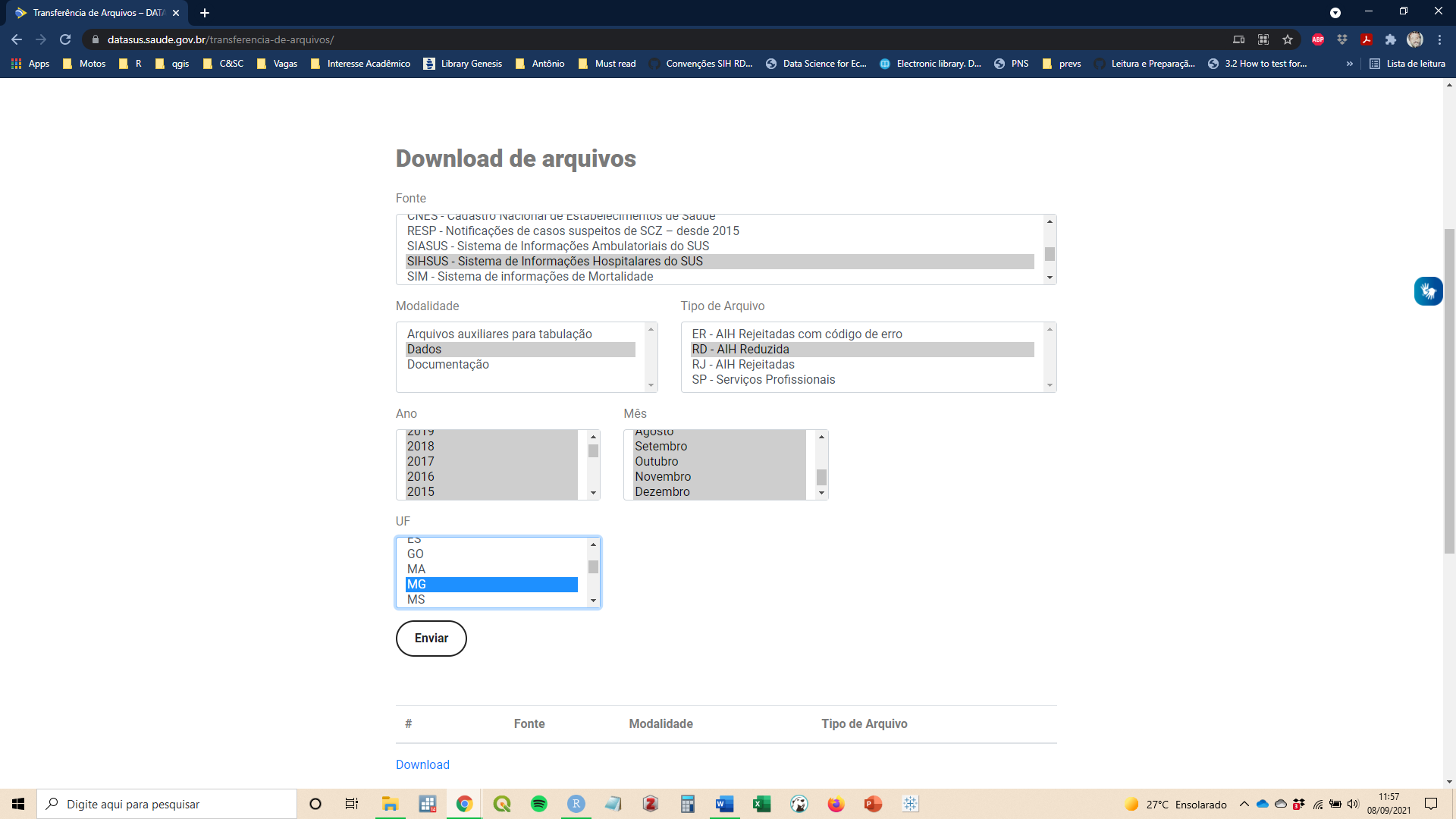

7. The selected files will be listed, andat the bottom of the list you will see the download button. Files are named as ‘DP’+name of the STATE (MG) +year(YY)’.dbc’

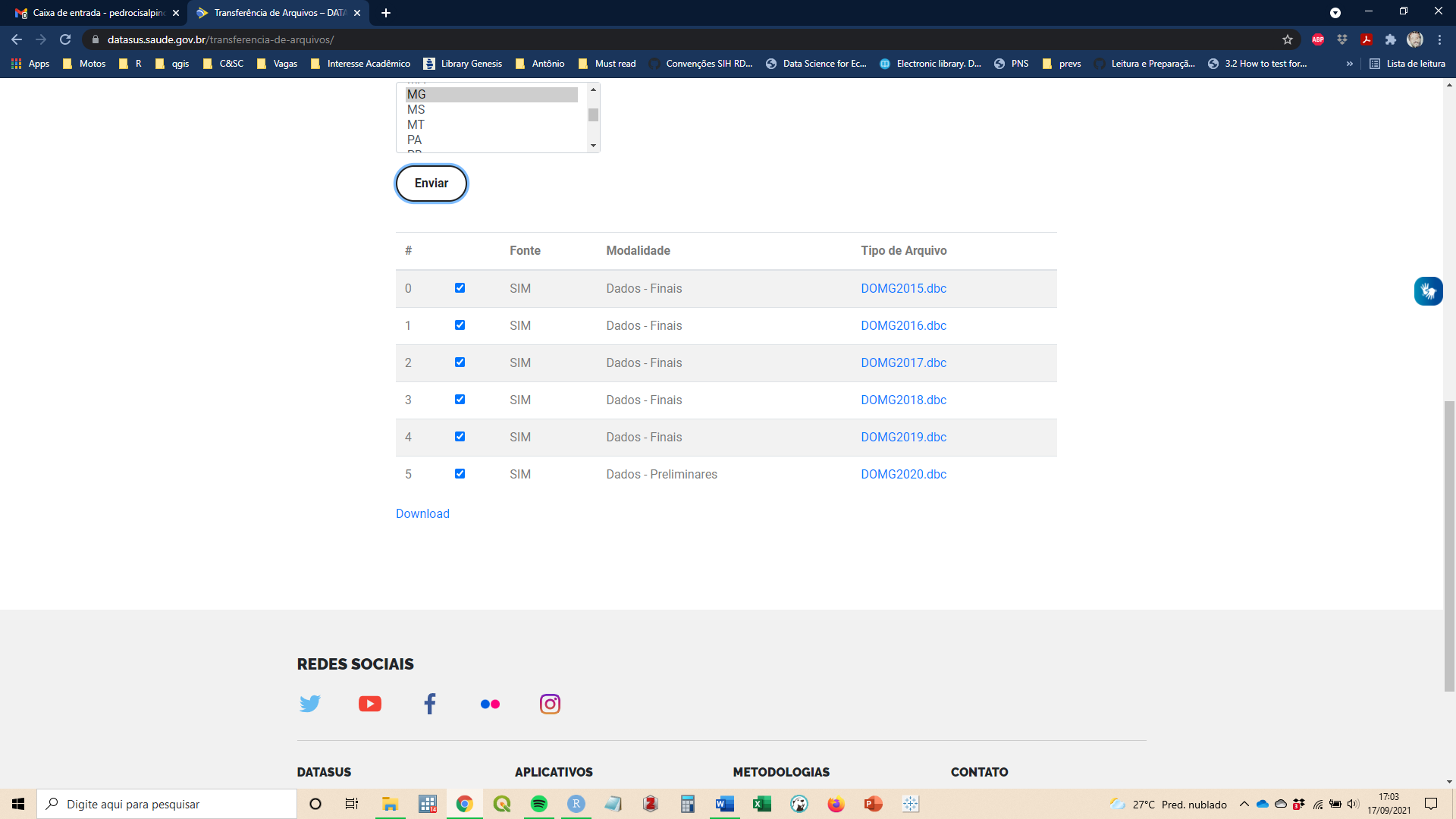
