## Supplementary material for "Cardiovascular Mortality during the COVID-19 Pandemics in a Large Brazilian City: a Comprehensive Analysis": Sup_Material_3

|  | Observed | Expected | Absolute Difference | Risk Ratio |
| --- | --- | --- | --- | --- |
| Deaths | 33  (30;37) | 39  (36;43) | -6.4 | 0.84  (0.73;0.96)* |
| Deaths at Home | 21  (19;24) | 25  (22;28) | -3.7 | 0.85  (0.72;1.01) |
| % Deaths at Home | 64  (59;69) | 63  (59;68) | 1.0 | 1.02  (0.86;1.2) |
| Hosp. Admissions | 100  (95;106) | 98  92;104) | 2.4 | 1.02  (0.94;1.11) |
| ICU | 20  (18;23) | 19  (16;21) | 1.7 | 1.09  (0.9;1.31) |
| % ICU | 20  (18;22) | 19  (17;21) | 1.2 | 1.06  (0.88;1.28) |
| In-Hospital Deaths | 7  (5;9) | 8  (6;9) | -0.7 | 0.91  (0.67;1.23) |
| % In-Hospital Deaths | 7  (5;8) | 8  (6;9) | -0.9 | 0.89  (0.66;1.2) |

**Supplemental Material 3: Age-standardized rates per 100,000 inhabitants and proportions for ICD Chapter XVIII outcomes observed in 2020, and expected (mean of 2015-2019), for epidemiological weeks 10-48, their absolute difference, and risk ratio. Belo Horizonte MG, Brazil.**

ICD: International Classification of Diseases, ICU: intensive care unit
