## Supplementary material for "Cardiovascular Mortality during the COVID-19 Pandemics in a Large Brazilian City: a Comprehensive Analysis": Sup_Material_4

| **Variable** | **30-59** | | | | **60+** | | | |
| --- | --- | --- | --- | --- | --- | --- | --- | --- |
|  | **Observed** | **Expected** | **Absolute Difference** | **Risk Ratio** | **Observed** | **Expected** | **Absolute Difference** | **Risk Ratio** |
| Deaths | 50  (46;55) | 55 (50;61) | -5.11 | 0.91 (0.79;1.04) | 734 (706;764) | 715  (686;744) | 19.63 | 1.03  (0.97;1.09) |
| Deaths at Home | 15  (13;18) | 14 (12;17) | 1.31 | 1.09 (0.85;1.4) | 253 (236;271) | 185  (170;200) | 68.20 | 1.37  (1.23;1.52)* |
| % Deaths at Home | 31  (26;35) | 25 (21;29) | 5.19 | 1.2 (0.93;1.54) | 34 (32;36) | 25  (24;27) | 8.59 | 1.33  (1.2;1.48)* |
| Hosp. Admissions | 387 (374;401) | 583 (567;600) | -196.07 | 0.66 (0.63;0.69)* | 1760 (1716;1807) | 2123  (2074;2174) | -362.80 | 0.83  (0.8;0.86)* |
| ICU | 150  (141;158) | 175 (166;184) | -25.15 | 0.86 (0.79;0.92)* | 669 (642;698) | 751  (722;782) | -82.43 | 0.89  (0.84;0.94)* |
| % ICU | 38  (37;40) | 30  (28;31) | 8.69 | 1.29 (1.19;1.39)* | 38 (36;39) | 35  (34;36) | 2.61 | 1.07  (1.01;1.14)* |
| In-Hospital Deaths | 14  (12;17) | 18  (15;21) | -3.77 | 0.8 (0.63;1.01) | 135 (123;149) | 148  (135;162) | -12.69 | 0.91  (0.8;1.04) |
| % In-Hospital Deaths | 3  (3;4) | 3  (2;3) | 0.64 | 1.2 (0.95;1.53) | 7  (7;8) | 7  (6;7) | 0.73 | 1.1  (0.97;1.25) |

**Supplemental Material 4: Age-standardized rates per 100,00 inhabitants and proportions for cardiovascular disease outcomes observed in 2020, and expected (mean of 2015-2019) for epidemiological weeks 10-48, their absolute difference, and risk ratio, according to age range.Belo Horizonte, MG, Brazil.**

ICU: intensive care unit
